# Prevalence of SARS-CoV-2 Infections in a Pediatric Orthopedic Hospital

**DOI:** 10.1101/2020.08.31.20183533

**Authors:** Ghalib Bardai, Jean A. Ouellet, Thomas Engelhardt, Gianluca Bertolizio, Zenghui Wu, Frank Rauch

## Abstract

This project assessed the prevalence of active and past infection with severe acute respiratory syndrome coronavirus 2 (SARS-CoV-2) in a specialized pediatric institution that did not provide care for coronavirus disease 2019 (Covid-19). The study was performed in Montreal, the city with the highest number of Covid-19 cases in Canada during the early phase of the pandemic. Testing for SARS-CoV-2 RNA in 199 individuals (39 children, 61 accompanying persons, 99 hospital employees) did not reveal active infection in any of the study participants. However, 22 (11%) of study participants had SARSCoV-2 IgG antibodies, indicating prior infection. Ten of these participants did not report symptoms compatible with Covid-19 in the 6 months prior to the study. Thus, although no evidence for active infection was found within the institution, consideration should be given to regular staff testing to detect asymptomatic spreading of SARS-CoV-2. In addition, it could be useful to test accompanying persons in children presenting for surgical procedures.

## Introduction

Montreal has the highest incidence of coronavirus disease 2019 (Covid-19) among cities in Canada, with 29,596 reported cases as of August 22, 2020, representing about one quarter of all reported cases in Canada (1). The present study was performed in a pediatric institution located in Montreal that provides elective-only specialized services for children with musculoskeletal conditions, such as pediatric orthopedics, rehabilitation and medical therapies. The institution does not provide care for infectious diseases; patients with known severe acute respiratory syndrome coronavirus 2 (SARS-CoV-2) infections are therefore treated at other institutions. At the start of the Covid-19 pandemic, a set of measures was implemented to minimize the risk of infection within the institution, such as screening each person entering the hospital, mandatory temperature checks at the entrance, wearing face masks within the building, reducing the number of clinic visits and testing for SARS-CoV-2 RNA (using nasopharyngeal swabs) in patients scheduled for surgery (Figure 1).

**Figure 1.**
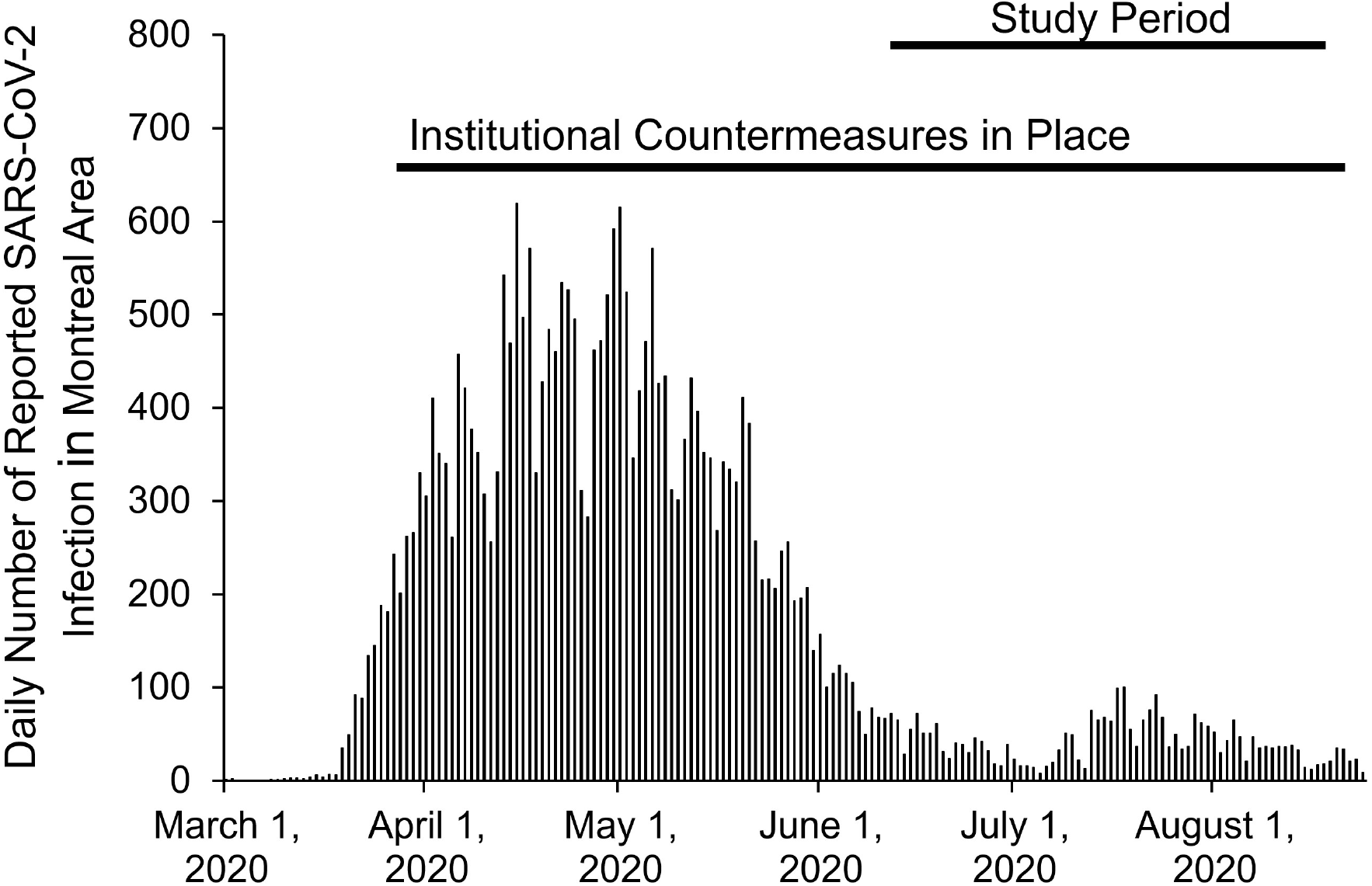
Study period in relation to infection rate in the Montreal area (data from: https://santemontreal.qc.ca). The timing of institutional countermeasures (i.e., wearing of face masks, mandatory temperature check at the hospital entrance; screening for Covid-19 related symptoms) is also indicated.

The aim of this project was to assess whether the institution remained free of SARS-CoV-2 infections after these countermeasures were implemented. To this end, patients, accompanying persons and employees were offered testing for active virus infection by examining for SARS-CoV-2 RNA in saliva. In addition, blood samples were obtained to test for SARS-CoV-2 IgG antibodies as indicators of past virus infection. As the sensitivity of detecting SARS-CoV-2 RNA in saliva samples was not well established at the start of the project, participating hospital employees underwent weekly sampling to increase the probability of identifying active infection with SARS-CoV-2.

## Subjects and Methods

Study participants were recruited at Shriners Hospital for Children – Canada in Montreal, Canada, between June 10 and July 27, 2020. All participants were asymptomatic for SARS-CoV-2 IgG antibodies infection at the time of recruitment, as only asymptomatic individuals had access to the hospital. The study population was comprised of 200 participants. This included 100 employees (about one third of the full-time staff) and 100 patients or individuals accompanying patients. All study participants or their legal guardians provided informed consent. The study was approved by the Institutional Review Board of McGill University (study number A05-M45-20B).

Study participants who were patients at the hospital or persons accompanying them had a single study visit. Hospital employees underwent 5 study visits on consecutive weeks. At each study visit, participants were asked to provide 2.5 ml of saliva into a sterile container and blood samples (5 ml) were obtained in serum tubes. Saliva samples were stored at -80C until analysis, serum was stored at 20C.

Saliva samples were tested for the presence of SARS-CoV-2 virus using real-time PCR (ABI 7500 Fast, Applied Biosystems). Viral nucleic acid isolation for SARS Cov-2 was performed using the Applied Biosystems MagMAX Viral/Pathogen kit on a Kingfisher (Thermo Fisher) instrument according to the manufacturers protocol. Extracted RNA was analysed using a commercially available kit (Taqpath RT-PCR COVID-19 Combo Kit with TaqPath 1-Step Multiplex Master Mix, Applied Biosystems). The method assesses for the presence of three SARS-CoV-2 target genes (Orf1a/b, N and S). Data analysis and interpretation of PCR data was performed using the Applied Biosystems COVID-19 Interpretive Software v1.2.

Serum samples were tested for the presence of SARS-CoV-2 IgG antibodies using a commercially available enzyme-linked immunosorbent assay (ELISA) targeting SARS-CoV-2 Spike (S2) and Nucleoprotein (N) (Mologic, Omega diagnostics, UK) following the manufacturer’s instructions. Each plate had a diluent (negative), cut off and positive controls. Results were considered positive if they were 10% above the cut off value. Extensive validation of this assay found a specificity of 97% (2). Group differences in categorical variables were tested for significance using the chi-square test. All tests were two-tailed and throughout the study P-values <0.05 were considered significant. Calculations were performed using SPSS Software Version 24 for Windows (SPSS, Inc., Chicago, IL, USA).

## Results

Results were obtained from 199 individuals (139 females, 60 males) (Table 1). One of the employees left the institution before any sample was obtained; therefore, results are available on 199 study participants. None of the study participants had detectable SARS-CoV-2 RNA in saliva samples, but 22 (11%) study participants were positive for SARS-CoV-2 IgG antibodies in serum samples. The difference in seroprevalence between patients, accompanying persons and hospital employees was not statistically significant (P = 0.75).

**Table 1.** Demographics of study population and study results.

|  | Patients | Accompanying<br>Persons | Employees |
| --- | --- | --- | --- |
| N | 39 | 61 | 99 |
| Females, N (%) | 24 (62%) | 41 (67%) | 74 (75%) |
| Age (years) | 15.6 (13.4; 16.8) | 47.1 (41.4; 50.8) | 42.5 (32.5; 52.5) |
| SARS-CoV-2 positive by PCR, N (%) | 0 (0%) | 0 (0%) | 0 (0%) |
| SARS-CoV-2 IgG antibody positive, N (%) | 3 (8%) | 7 (12%) | 12 (12%) |

Results are shown in N (%) or as median (interquartile range).

Of the 22 seropositive individuals, 12 reported symptoms compatible with Covid-19 in the 6 months prior to the study. Thus, 10 seropositive study participants (1 child, 3 accompanying persons, 6 employees) were considered asymptomatic. Eleven (50%) seropositive individuals had undergone prior SARS-CoV-2 testing by nasopharyngeal swab 4 or more weeks prior to the present study, which had yielded a positive result in 5 of these study participants. Four individuals (3 employees, 1 accompanying person) had provided care to patients with Covid-19 infections at other health care institutions, the other seropositive study participants had no known exposure to the virus.

Among hospital employees, SARS-CoV-2 IgG antibodies were found in 3 of 20 (15%) operation room personnel, in 5 of 44 (11%) health care workers from other departments and in 4 of 35 (11%) employees who did not have direct patient contact (P = 0.91). Eleven employees were seropositive at the first study visit. One employee had SARS-CoV-2 antibodies only at the last study visit, even though no SARS-CoV-2 RNA had been found in any of the employee’s saliva samples. Among the 12 employees with SARS-CoV-2 IgG antibodies in at least one sample, 3 (25%) had a negative antibody test result by the time of their last study visit.

## Discussion

In this study we found that none of the study participants had detectable active infection with SARSCoV-2 virus at the time of the study. A prior study had found that use of saliva samples was a sensitive method to detect SARS-CoV-2 in asymptomatic health care workers (3). However, 11% of study participants had IgG antibodies against SARS-CoV-2, indicating prior infection with the virus. Seroprevalence was similar between hospital employees, patients and accompanying persons and did not vary with the type of work performed by employees.

The prevalence of SARS-CoV-2 IgG antibodies among hospital employees in the present study is similar to the observations in a New York hospital system, where 13.7% of health care workers were seropositive (4). Our findings are also in line with a report from a large Belgian tertiary health care center, where seroprevalence was similar among employees who had patient contact and those without patient contact (5). However, the overall seroprevalence of 11% in the present study is higher than what has been reported in a study on blood donors in Montreal that was conducted at approximately the same time as the present study (between May 25 and July 9), where only 3% were seropositive (6). Even though this study used a different IgG antibody than the present study, it appears that the SARSCoV-2 seroprevalence is higher among individuals who volunteer for participation in a SARS-CoV-2 study than among blood donors.

While 12 study participants with SARS-CoV-2 antibodies reported symptoms compatible with Covid19 infection in the 6 months prior to the study, 10 individuals, including 6 employees, appeared to have had asymptomatic infection. The fact that 6 of the 99 participating employees had undergone apparently asymptomatic infection suggests that testing of asymptomatic employees for virus RNA would have been necessary to identify their SARS-CoV-2 infection. Nevertheless, the observation that one employee developed antibodies despite repeatedly negative saliva tests shows that even repeated testing may not correctly identify all individuals with SARS-CoV-2 infection.

The finding that 3 of the 12 seropositive study participants had a negative SARS-CoV-2 antibody result at the last study visit is in accordance with previous reports that in up to 40% of those recovering from mild disease, SARS-CoV-2 antibodies fall to undetectable levels within 2-3 months (7). However, recent data suggest that antibodies against the N protein decline faster than antibodies against the S2 protein (8). The ELISA used in the present study used antibodies directed against both of these proteins and may therefore reflect the decline in N protein specific antibodies.

Thus, while no case of active SARS-CoV-2 infection was detected among individuals within our health care institution there was evidence for prior infection in 11% of study participants. Consideration should be given to regular staff testing to detect asymptomatic spreading of SARS-CoV-2. In addition, it could be useful to test accompanying persons in children presenting for surgical procedures.

## Data Availability

The data that support the findings of this study are available from the corresponding author upon reasonable request.

## Acknowledgements

We thank Michaela Durigova, Marie-Josée Giguère and Elizabeth Parr for organizational support, Vanessa Pépin, Carine Dahan, Carla Evangeliste, Souad Rhalmi and Juliana Marulanda for sample procurement, and Charlotte El-Mir, Sami Abdullah and Faviola Huapaya-Wong for sample analysis. The study was supported by the Shriners of North America. ELISA IgG antibody plates were donated by Mologic.

## Notes

### Competing Interest Statement

The authors have declared no competing interest.

### Clinical Trial

This is an observational study. No intervention was performed.

### Funding Statement

The study was supported by the Shriners of North America. ELISA IgG antibody plates were donated by Mologic Inc.

### Author Declarations

Institutional Review Board of McGill University (study number A05-M45-20B)

